# Associations of Serum Folate and Vitamin B_12_ levels with Cardiovascular Mortality among Non-diabetic Population

**DOI:** 10.1101/2025.01.13.25320498

**Authors:** Zijun Chen, Dian Cheng, Xuecheng Wang

**Author notes:** Corresponding author: Xuecheng Wang, Department of Cardiovascular Medicine, Shanghai East Hospital, School of Medicine, Tongji University, 150 Jimo Road, Shanghai 200120, P.R. China. Zijun Chen and Dian Cheng, contributed equally to this work and should be considered co-first authors.

## Abstract

**Background:** While studies have examined the associations between folate, vitamin B_12_, and cardiovascular mortality in high-risk populations, evidence regarding these associations in non-diabetic population remains limited. This study aimed to assess the associations of serum folate and vitamin B_12_ levels with cardiovascular mortality in a large, nationally representative cohort of US non-diabetic adults.

**Methods and Results:** Data were extracted from the National Health and Nutrition Examination Survey (NHANES) cycles from 1999 to 2006 and 2011 to 2014. Multivariate logistic regression and restricted cubic spline (RCS) analyses were used to evaluate the associations between serum folate, vitamin B_12_ and cardiovascular mortality. A two-piecewise logistic regression model was employed to identify the inflection points of serum folate and vitamin B_12_ levels on cardiovascular mortality. Among 19,402 US adults, the mean age was 43.9 ± 18.6 years, 9,749 (50.2%) were male, and 8,438 (43.5%) were non-Hispanic White. With a median follow-up of 159 months (IQR: 82.0-193.0 months), 928 cardiovascular deaths were documented. After fully adjusting for potential confounders, a significant inverse association was observed between serum folate levels and cardiovascular mortality. The RCS analysis revealed non-linear associations between serum folate (*P* for nonlinearity = 0.02), vitamin B_12_ levels (*P* for nonlinearity = 0.04), and the risk of cardiovascular mortality. Specifically, serum folate levels below 19.50 ng/mL and serum vitamin B_12_ levels at or above 436.52 pg/mL were both linked to an increased risk of cardiovascular mortality. Stratified and sensitivity analyses confirmed the robustness of these associations.

**Conclusions:** This study demonstrated that low serum folate levels and high vitamin B_12_ levels were associated with an increased risk of cardiovascular mortality in non-diabetic individuals. These findings provide evidence supporting the potential of serum folate and vitamin B_12_ levels as biomarkers for cardiovascular mortality risk estimation, pending further validation in other independent cohorts.

## 1. Introduction

Cardiovascular disease (CVD) remains the leading cause of mortality worldwide.^1^ In the United States, an estimated 127.9 million adults (48.6%) are affected by CVD, including coronary heart disease (CHD), heart failure, hypertension, and stroke. Among individuals under 85, CVD accounts for more deaths than cancer, imposing substantial health and economic burdens both nationally and globally.^2^

Folate and vitamin B_12_ are essential water-soluble vitamins that play critical roles in one-carbon and homocysteine metabolism.^3,4^ Hyperhomocysteinemia (HHcy) is associated with increased CVD risk and mortality.^5,6^ Since folate and vitamin B_12_ can lower plasma homocysteine levels, they are considered potentially beneficial in the prevention and management of CVD.^7,8^ However, previous studies on the associations between folate, vitamin B_12_, and cardiovascular mortality have yielded inconsistent findings.

In general population studies, some reported that dietary folate intake is inversely associated with cardiovascular mortality, while no association was found between vitamin B_12_ intake and mortality risk.^9^ Other studies have indicated that elevated red blood cell folate concentrations may be associated with a higher risk of CHD.^10^ Additionally, an observational study in diabetic population showed that both low and high serum vitamin B_12_ levels, as well as low serum folate levels, are significantly associated with cardiovascular mortality.^11^ Despite this, limited research has specifically investigated the associations between folate, vitamin B_12_, and cardiovascular mortality in non-diabetic population. In these individuals, risk factors such as hypertension, hypercholesterolemia, smoking, physical inactivity, and obesity also significantly elevate CVD incidence.^12^ Thus, further studies across various populations are essential to provide more evidence.

This study aims to clarify the associations of serum folate and vitamin B_12_ levels with cardiovascular mortality among non-diabetic population.

## 2. Methods

### 2.1 Data source and study population

The data for this study were obtained from the National Health and Nutrition Examination Survey (NHANES), a nationally representative, cross-sectional survey employing a stratified, multistage probability sampling design. Conducted by the National Center for Health Statistics (NCHS) of the Centers for Disease Control and Prevention (CDC), NHANES evaluates the health and nutritional status of the general US population. The survey methodology and data collection procedures have been previously documented and published.^13^ The NHANES research protocol received approval from the NCHS Ethics Review Board, and all participants provided written informed consent upon enrollment. All data analyzed in this study are publicly available online (https://wwwn.cdc.gov/nchs/nhanes/Default.aspx).

For this cohort study, we used data from six NHANES cycles (1999-2000, 2001-2002, 2003-2004, 2005-2006, 2011-2012, and 2013-2014), selected based on the availability of both serum folate and vitamin B_12_ measurements. Notably, serum vitamin B_12_ data were unavailable for the NHANES cycles from 2007-2010. The non-diabetic population included individuals with normoglycemia and prediabetes. Normoglycemia was defined as fasting blood glucose (FBG) *<* 100 mg/dL and glycosylated hemoglobin A1c (HbA1c) *<* 5.7%, while prediabetes was defined by FBG between 100 and 125 mg/dL or HbA1c between 5.7% and 6.4%.^14^ Exclusion criteria were as follows: (1) Diabetic population (n = 4,686): defined by self-reported diagnosis of diabetes, use of anti-diabetic medications or insulin, HbA1c ≥ 6.5%, FBG ≥ 126 mg/dL, or 2-hour postprandial glucose ≥ 200 mg/dL.^14^ (2) Participants aged *<* 18 years (n = 26,731); (3) Pregnant participants (n = 1,095); (4) Participants with a history of cancer or malignancy (n = 2,203); (5) Participants missing serum folate and Vitamin B_12_ measurements (n = 7,263); (6) Participants without eligible follow-up data (n = 25). Finally, a total of 19,402 non-diabetic participants were included in our analysis (**Figure 1**).

**Figure 1.**
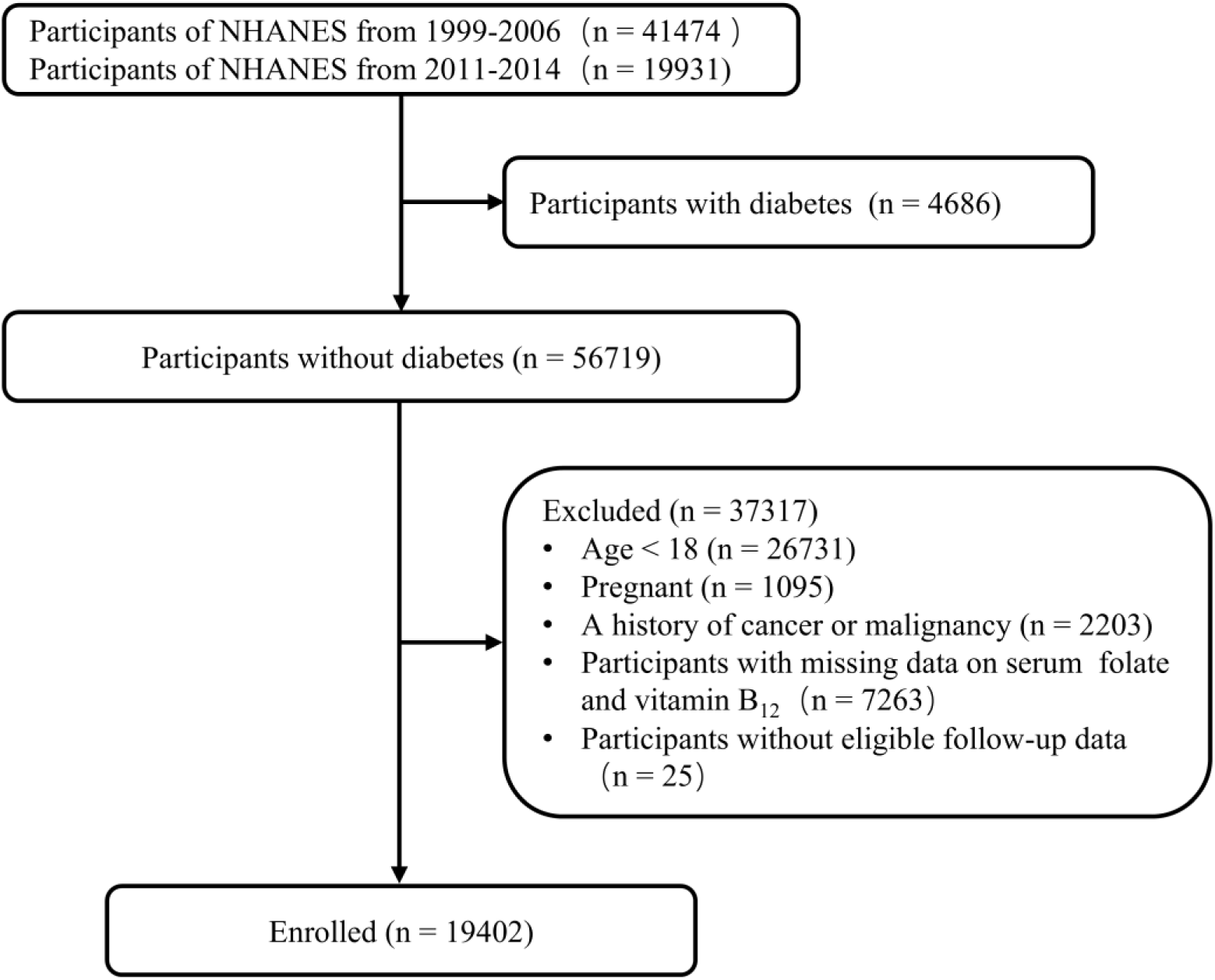
Flowchart of study participants. NHANES, National Health and Nutrition Examination Survey.

### 2.2 Measurements of serum folate and vitamin B_12_

Serum and whole blood samples were collected through venipuncture and sent to the Nutritional Biomarkers Laboratory at the CDC for analysis. Detailed descriptions of blood sample collection and processing are available on the NHANES website (https://www.cdc.gov/nchs/nhanes). This study utilized serum total folate and vitamin B_12_ levels. From 1999 to 2006, serum folate and vitamin B_12_ levels were measured using the Quantaphase II radioimmunoassay by Bio-Rad Laboratories. From 2011 to 2014, serum folate levels were measured using isotope-dilution high-performance liquid chromatography (HPLC) coupled with tandem mass spectrometry (MS/MS), while serum vitamin B_12_ levels were determined using the Roche electrochemiluminescence immunoassay. According to NHANES guidelines, serum folate and vitamin B_12_ levels from 1999-2006 were converted to equivalent values for 2007-2014 to correct for systematic bias and ensure data comparability across different periods.

### 2.3 Outcome definition

To ascertain the mortality status of the follow-up participants in this study, we utilized the NHANES public-use linked mortality file through December 31, 2019. This file was linked to the National Death Index (NDI) database using a probabilistic matching algorithm, which is widely regarded as a highly reliable source for identifying mortality.^15^ The specific causes of death were determined according to the International Classification of Diseases, Tenth Revision (ICD-10). Cardiovascular mortality includes any death related to cardiovascular disease, encompassing codes and I00-I09, I10, I11, I13, and I20-I51.

### 2.4 Covariates

In this study, the covariates included demographic data, lifestyle factors, medical history, physical measurements, laboratory tests, and NHANES survey cycles. Demographic, lifestyle, and medical history information were collected through standardized interview questionnaires administered at Mobile Examination Centers (MECs). Physical measurements and laboratory tests were conducted according to standardized protocols. The detailed acquisition process and testing methods are available at http://www.cdc.gov/nchs/nhanes/. Demographic data included age (< 65, ≥ 65), gender (male, female), race (non-Hispanic White, non-Hispanic Black, Mexican American, others), education level (below high school, high school, and above), and poverty-to-income ratio (PIR) (< 1, 1-3, > 3). Lifestyle information encompassed smoking status, alcohol consumption, physical activity, and the healthy eating index-2015 (HEI-2015). Smoking status was categorized into never, ever and current based on whether participants had smoked fewer than 100 cigarettes in their lifetime and the current smoking habits. Alcohol consumption was assessed with the question, “Have you ever had at least 12 drinks in your lifetime?” Responses were categorized as “Yes” or “No”. The HEI-2015 score indicated overall dietary quality, with higher scores reflecting better dietary quality.^16^ Physical activity was classified into low (< 600 min/week), moderate (600 - 1200 min/week), and high levels (≥ 1200 min/week).^17^ Medical history included hypertension, hypercholesterolemia, heart failure, CHD, angina, acute myocardial infarction (AMI), and stroke. Hypertension was defined as a self-reported diagnosis of hypertension, systolic blood pressure (SBP) > 140 mmHg and/or diastolic blood pressure (DBP) ≥ 90 mmHg, or currently receiving antihypertensive medication. Hypercholesterolemia was defined as a self-reported hypercholesterolemia, total cholesterol level ≥ 200 mg/dL, or use of cholesterol-lowering medication. A history of heart failure, CHD, angina, AMI, or stroke was determined through self-reported questionnaires. Physical measurements included SBP, DBP, height, and weight. Body mass index (BMI) was calculated by weight (kg) / height (m²) and classified as normal (< 25 kg/m²), overweight (25 - 29.9 kg/m²), and obese (≥ 30 kg/m²). Laboratory tests included triglycerides (TG), total cholesterol (TC), low-density lipoprotein cholesterol (LDL-C), high-density lipoprotein cholesterol (HDL-C). The estimated glomerular filtration rate (eGFR) was calculated using the Chronic Kidney Disease Epidemiology Collaboration (CKD-EPI) equation.^18^

### 2.5 Statistical analysis

Given the complex, multistage sampling design of the NHANES, all analyses in this study were performed in accordance with NHANES analytical guidelines, utilizing sampling weights, clustering, and stratification methods. For baseline characteristics, mean (standard deviation, SD) or median (interquartile range, IQR) was used to describe continuous variables, while counts (percentage, %) were used for categorical variables. Missing data were imputed using the ‘mice’ package, which applies a random forest algorithm for multiple imputation. Serum folate and vitamin B_12_ concentrations were log_10_-transformed and divided into quartiles, with the lowest quartile defined as the reference group. One-way ANOVA and Chi-square tests were used to compare serum folate and vitamin B_12_ levels across groups for continuous and categorical variables, respectively.

Multivariate logistic regression models were employed to estimate hazard ratios (HRs) and 95% confidence intervals (CIs) for associations of serum folate and vitamin B_12_ levels with cardiovascular motality. Three models were gradually applied. Model 1 adjusted for age, gender, and race. Model 2 adjusted for education level, PIR, smoking status, alcohol consumption, physical activity, and HEI-2015, based on Model 1. Glycemic status, hypertension, hypercholesterolemia, hypertension medication use, hyperlipidemia medication use, heart failure, CHD, angina, AMI, stroke, BMI, eGFR, TG, TC, LDL-C, HDL-C, and NHANES cycles were further adjusted in Model 3.

To investigate the non-linear relationships of serum folate and vitamin B_12_ levels with cardiovascular mortality, a restricted cubic spline (RCS) analysis with 4 knots (at the 5th, 35th, 65th and 95th percentiles) was performed within the range from the 5th to the 95th percentiles to minimize the influence of potential outliers. If nonlinearity was detected, we then applied a two-piecewise logistic regression model to examine the threshold effect of the log_10_-transformed serum folate and vitamin B_12_ levels on cardiovascular mortality, aiming to identify the inflection point. Additionally, a log-likelihood ratio test was conducted to compare the two-piecewise logistic regression model with a standard logistic regression model.

Stratified analyses were performed to assess the potential effect modification across age (< 65, or ≥ 65), gender (male, or female), race (non-Hispanic White, or other races), education level (below high school, or high school and above), PIR (<1, or ≥ 1), smoking status (yes, or no), alcohol consumption (yes, or no), BMI (< 25, or ≥ 25 kg/m²), HEI-2015 score (low, or high), physical activity (low, or high), CVD (history of heart failure, CHD, angina, and AMI) (yes, or no), glycemic status (normoglycemia, or prediabetes), hypertension (yes, or no), hypercholesterolemia (yes, or no) and eGFR (< 90, or ≥ 90 mL/min/1.73m^2^). Interaction tests were used to assess the differential effects across subgroups in the multivariate logistic regression model.

We also conducted sensitivity analyses to evaluate the robustness of the results.

(1) Adults with a history of CVD (heart failure, CHD, angina, and AMI) were excluded from the main analyses. (2) Adults who died within the first 2 years after participation were not included. (3) We further adjusted for homocysteine levels.

All statistical analyses were conducted using R software (version 3.6.0). A two-tailed *P*-value of < 0.05 was considered statistically significant.

## 3. Results

### 3.1 Participants characteristics

The baseline characteristics of participants across quartiles of serum folate and vitamin B_12_ levels were summarized in **Table 1**. Among 19,402 participants, the mean age was 43.9 ± 18.6 years, 9,749 (50.2%) were male, and 8,438 (43.5%) were non-Hispanic White. During a median follow-up of 159 months (IQR: 82.0-193.0 months), 928 cardiovascular deaths were documented. Compared to participants in the lowest quartile of serum folate levels (n = 4,872), those in the highest quartile (n = 4,807) were more likely to be older, female, never-smokers, with a higher education level, PIR, lower physical activity, lower BMI, and had a higher prevalence of prediabetes, hypertension, hypercholesterolemia, CVD and stroke. Similarly, individuals in the highest quartile of serum vitamin B_12_ levels (n = 4,847) were more likely to be older, female, never smokers, with a higher PIR, lower BMI, and had a higher prevalence of hypercholesterolemia compared to those in the lowest quartile (n = 4,857).

**Table 1.**
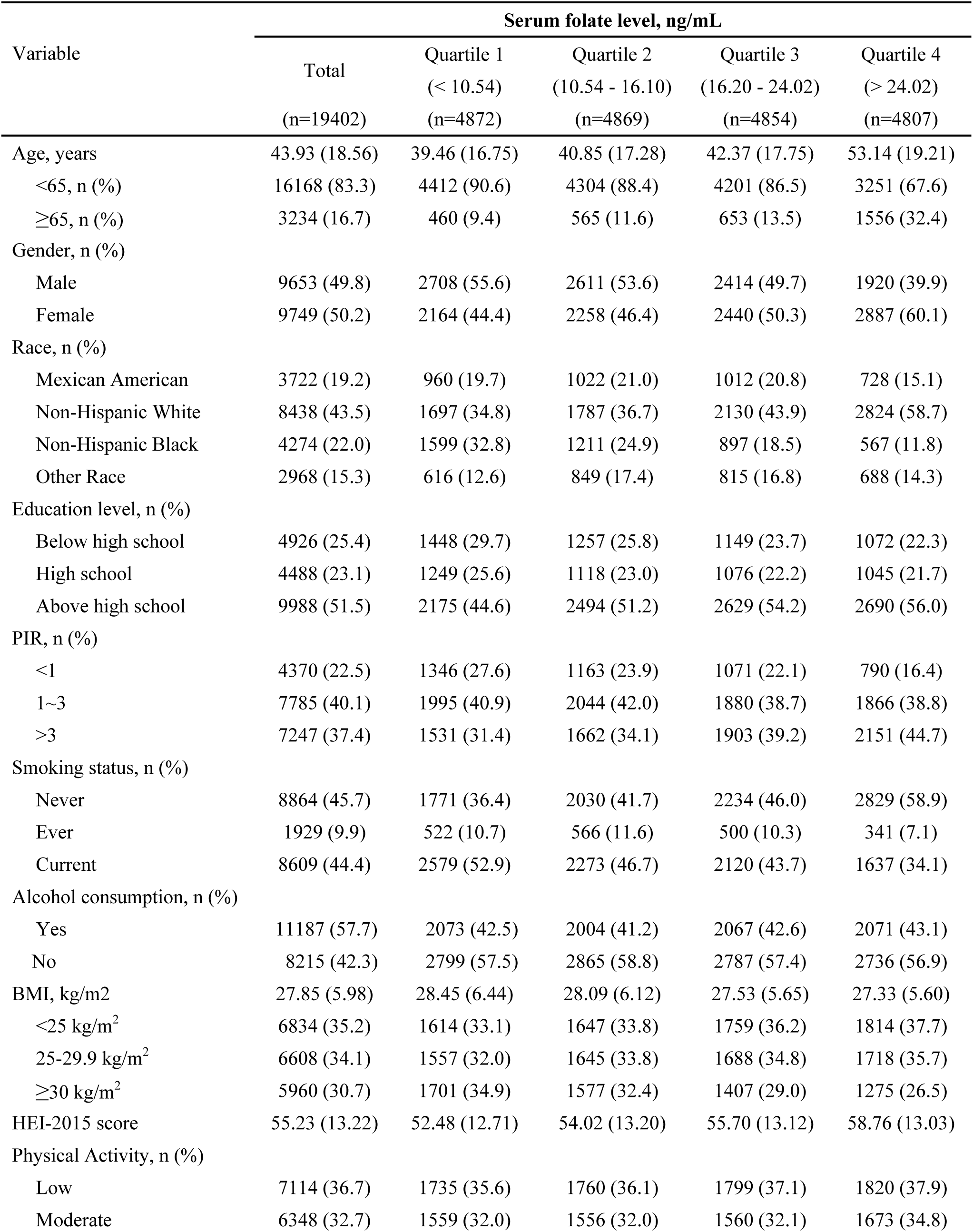
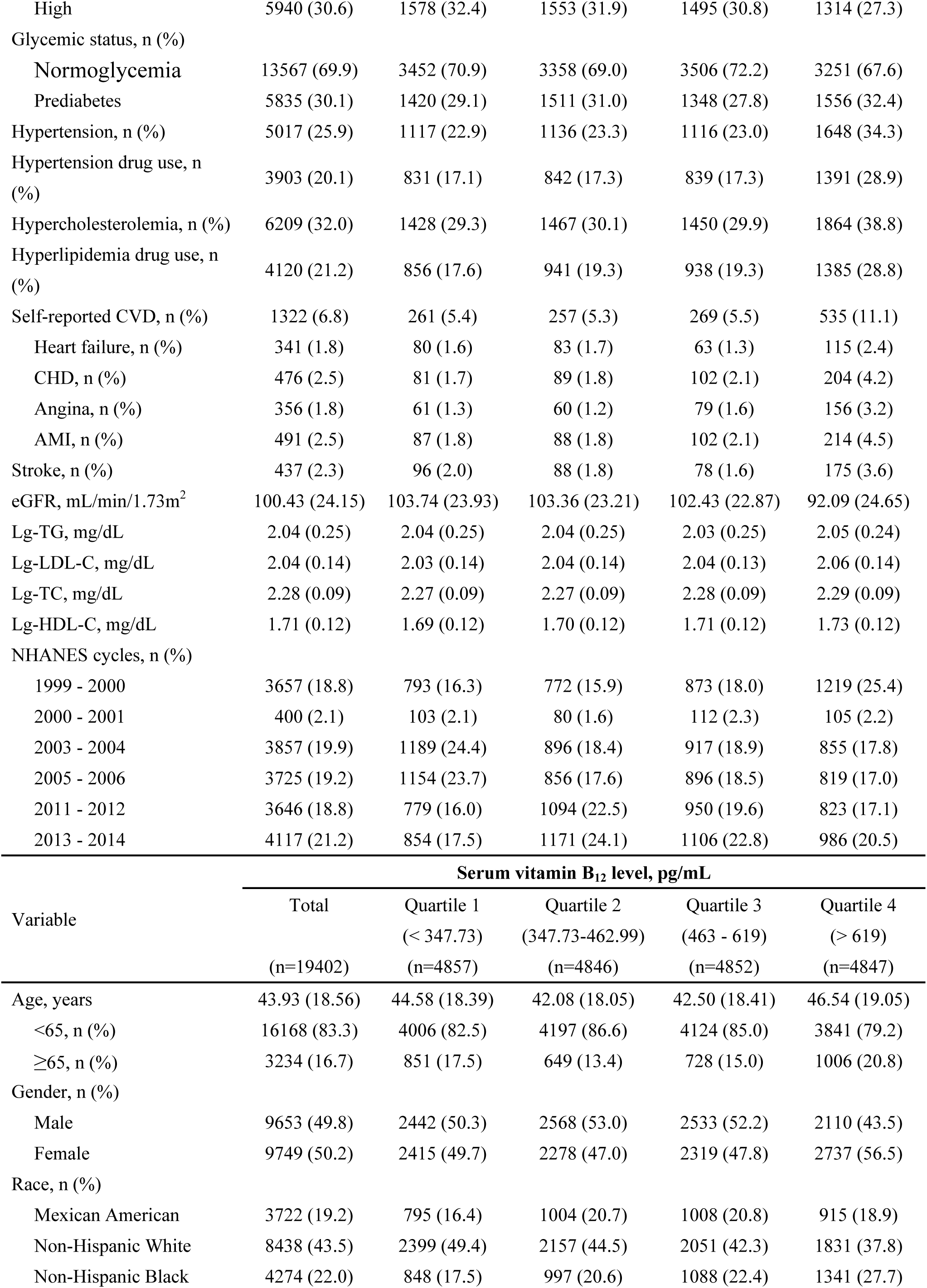
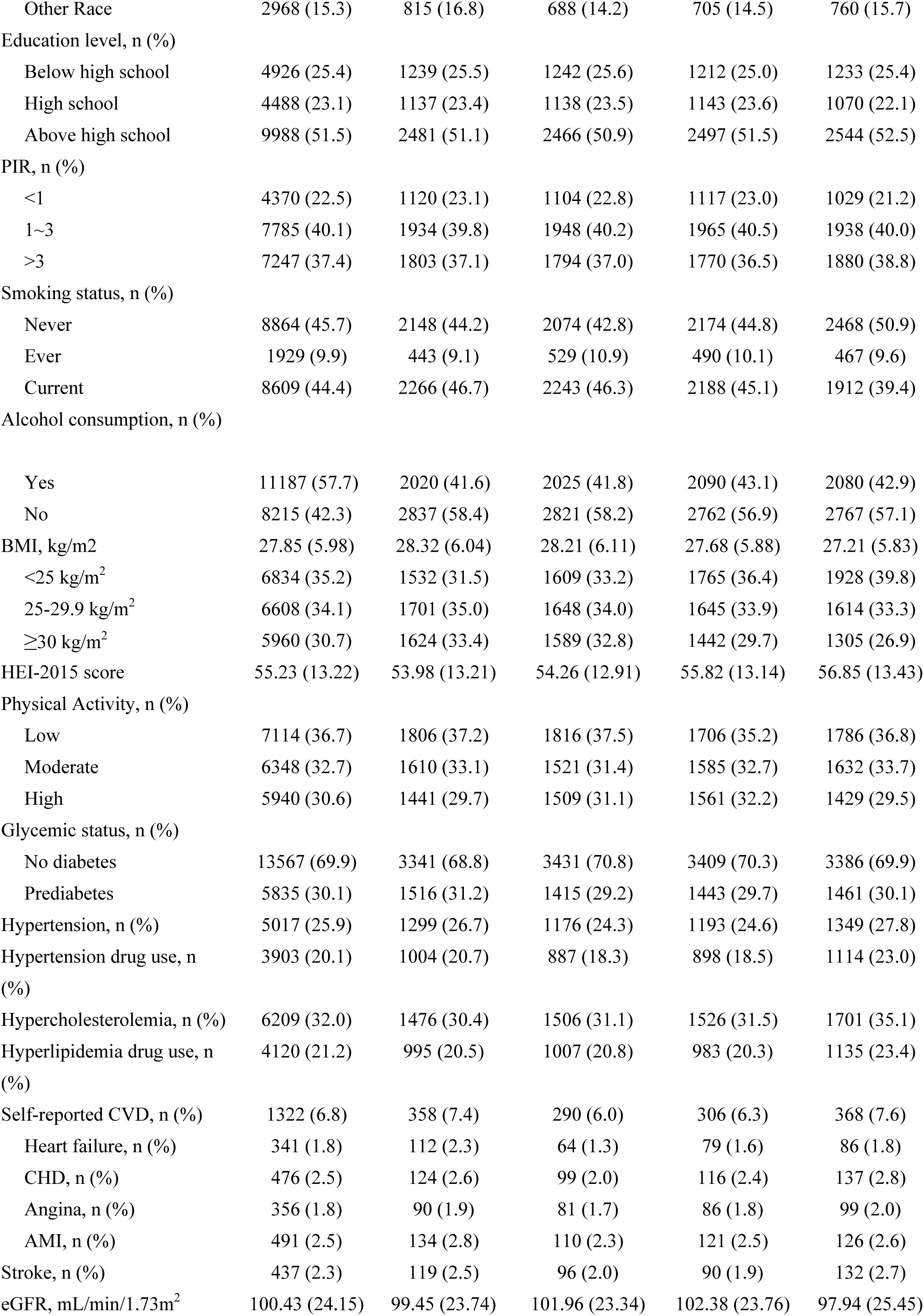
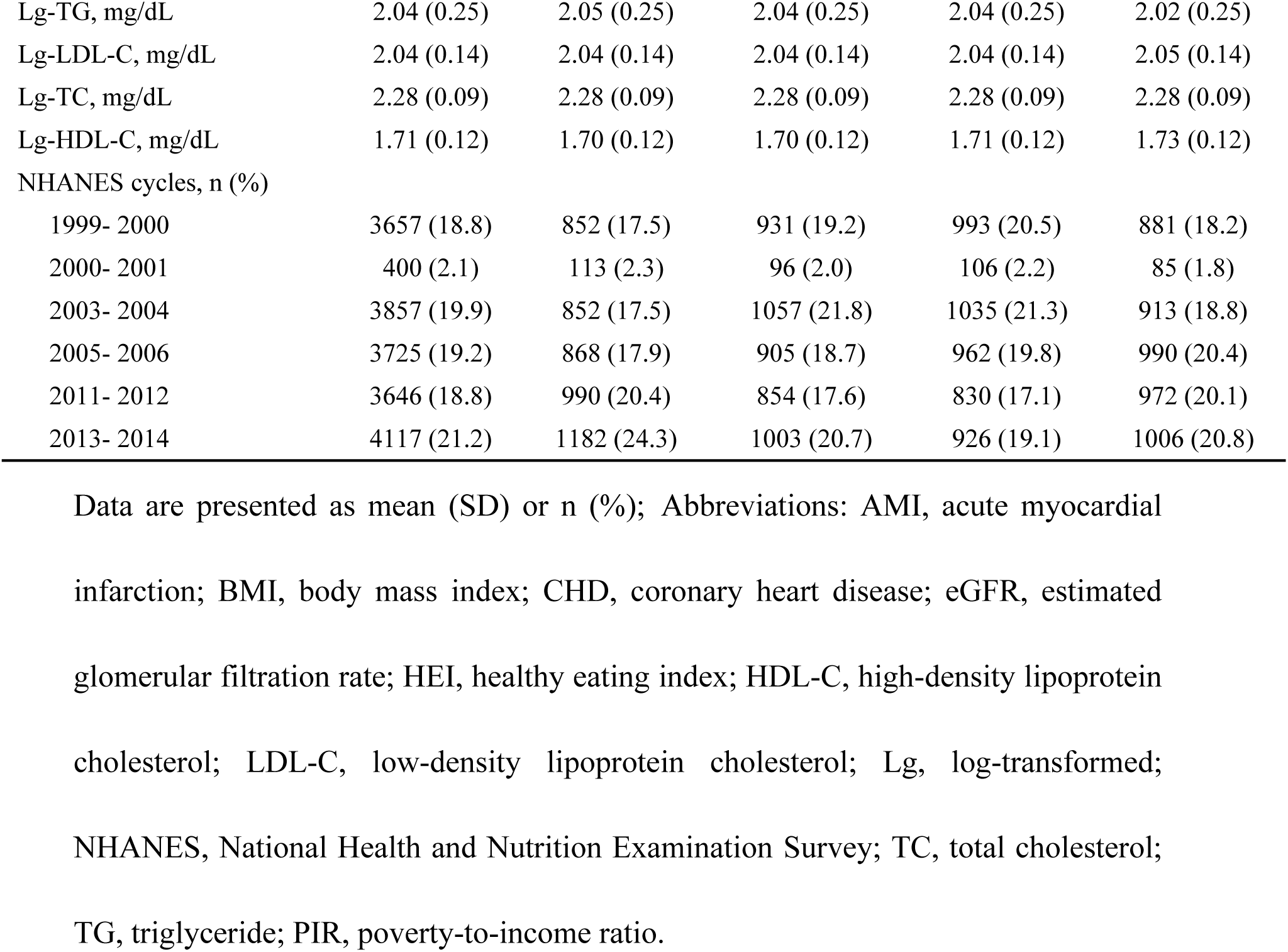
Baseline characteristics of participants with non-diabetes by serum folate and vitamin B_12_ levels.

### 3.2 Serum folate level and cardiovascular mortality

**Table 2** presents the associations of serum folate levels (in quartiles) with cardiovascular mortality. An inverse association was observed between serum folate levels and cardiovascular mortality in Models 1 and 2. After fully adjusting for potential confounders in Model 3, serum folate levels remained significantly associated with a lower risk of cardiovascular mortality. Specifically, compared with the reference group (the first quartile), the HRs of cardiovascular mortality were 0.67 (95% CI, 0.50-0.88) for the second quartile, 0.69 (95% CI, 0.53-0.88) for the third quartile, and 0.72 (95% CI, 0.56-0.91) for the fourth quartile.

**Table 2.**
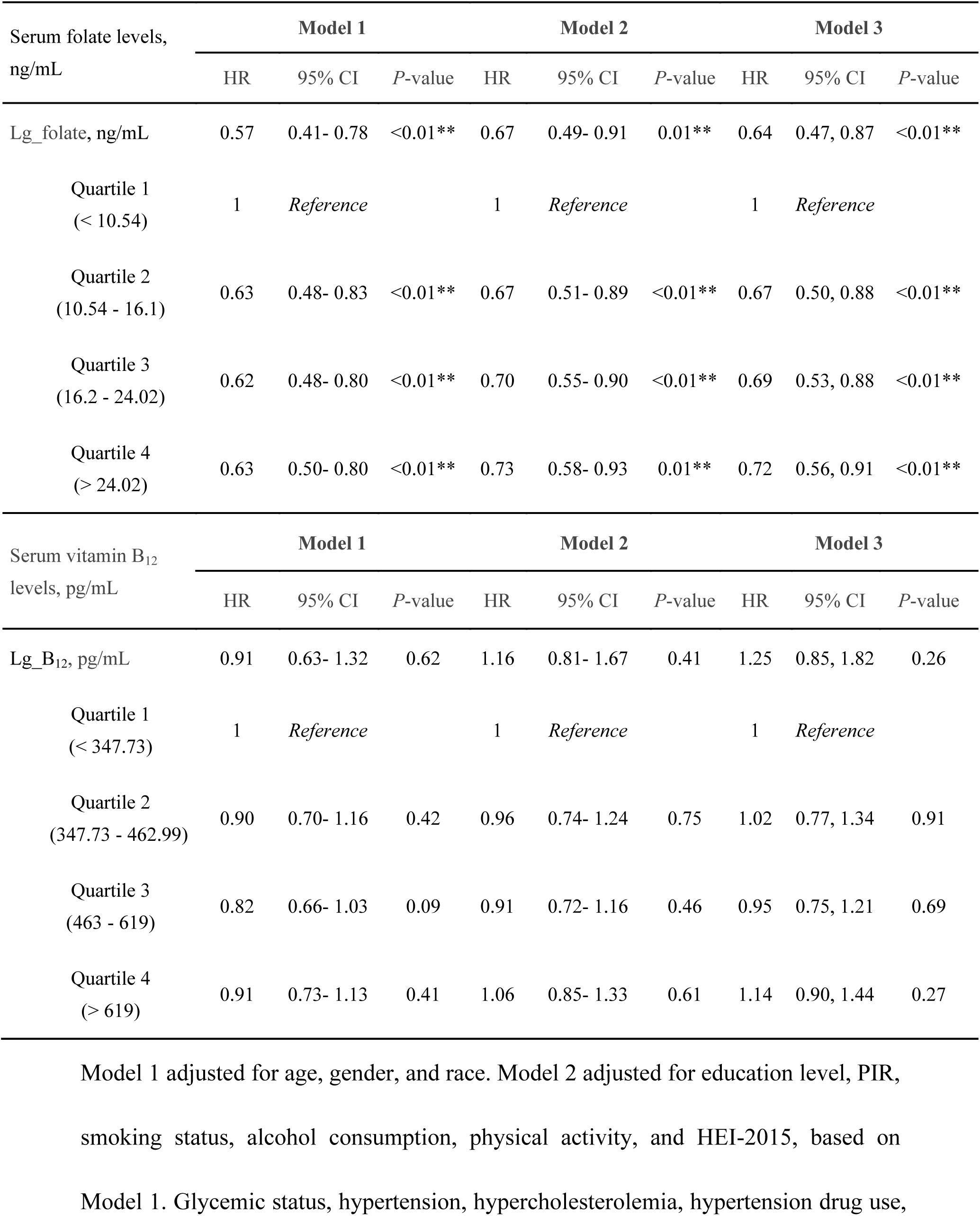
Hazard ratios of cardiovascular mortality by serum folate and vitamin B_12_ among adults with non-diabetes.

| Serum folate levels,<br>ng/mL | Model 1 |  |  | Model 2 |  |  | Model 3 |  |  |
| --- | --- | --- | --- | --- | --- | --- | --- | --- | --- |
|  | HR | 95% CI | P-value | HR | 95% CI | P-value | HR | 95% CI | P-value |
| Lg_folate, ng/mL | 0.57 | 0.41- 0.78 | <0.01** | 0.67 | 0.49- 0.91 | 0.01** | 0.64 | 0.47, 0.87 | <0.01** |
| Quartile 1<br>(< 10.54) | 1 | *Reference* |  | 1 | *Reference* |  | 1 | *Reference* |  |
| Quartile 2<br>(10.54 - 16.1) | 0.63 | 0.48- 0.83 | <0.01** | 0.67 | 0.51- 0.89 | <0.01** | 0.67 | 0.50, 0.88 | <0.01** |
| Quartile 3<br>(16.2 - 24.02) | 0.62 | 0.48- 0.80 | <0.01** | 0.70 | 0.55- 0.90 | <0.01** | 0.69 | 0.53, 0.88 | <0.01** |
| Quartile 4<br>(> 24.02) | 0.63 | 0.50- 0.80 | <0.01** | 0.73 | 0.58- 0.93 | 0.01** | 0.72 | 0.56, 0.91 | <0.01** |
| Serum vitamin B<sub>12</sub><br>levels, pg/mL | Model 1 |  |  | Model 2 |  |  | Model 3 |  |  |
|  | HR | 95% CI | P-value | HR | 95% CI | P-value | HR | 95% CI | P-value |
| Lg_B<sub>12</sub>, pg/mL | 0.91 | 0.63- 1.32 | 0.62 | 1.16 | 0.81- 1.67 | 0.41 | 1.25 | 0.85, 1.82 | 0.26 |
| Quartile 1<br>(< 347.73) | 1 | *Reference* |  | 1 | *Reference* |  | 1 | *Reference* |  |
| Quartile 2<br>(347.73 - 462.99) | 0.90 | 0.70- 1.16 | 0.42 | 0.96 | 0.74- 1.24 | 0.75 | 1.02 | 0.77, 1.34 | 0.91 |
| Quartile 3<br>(463 - 619) | 0.82 | 0.66- 1.03 | 0.09 | 0.91 | 0.72- 1.16 | 0.46 | 0.95 | 0.75, 1.21 | 0.69 |
| Quartile 4<br>(> 619) | 0.91 | 0.73- 1.13 | 0.41 | 1.06 | 0.85- 1.33 | 0.61 | 1.14 | 0.90, 1.44 | 0.27 |

The RCS analysis indicated a significant non-linear association between serum folate level and cardiovascular mortality risk (**Figure 2A**, *P* for nonlinearity = 0.02). The two-piecewise logistic regression analysis further revealed that the inflection point of serum folate was 19.50 ng/mL. When serum folate level was < 19.50 ng/mL, each 1-unit decrease in serum folate level exhibited an 55% increase in the risk of cardiovascular mortality (HR, 0.45; 95% CI, 0.29-0.70, *P* for log-likelihood ratio < 0.01). However, when serum folate level was ≥ 19.50 ng/mL, no significant association with cardiovascular mortality was observed (HR, 0.85; 95% CI, 0.38-1.88) (**Table 3**).

**Figure 2.**
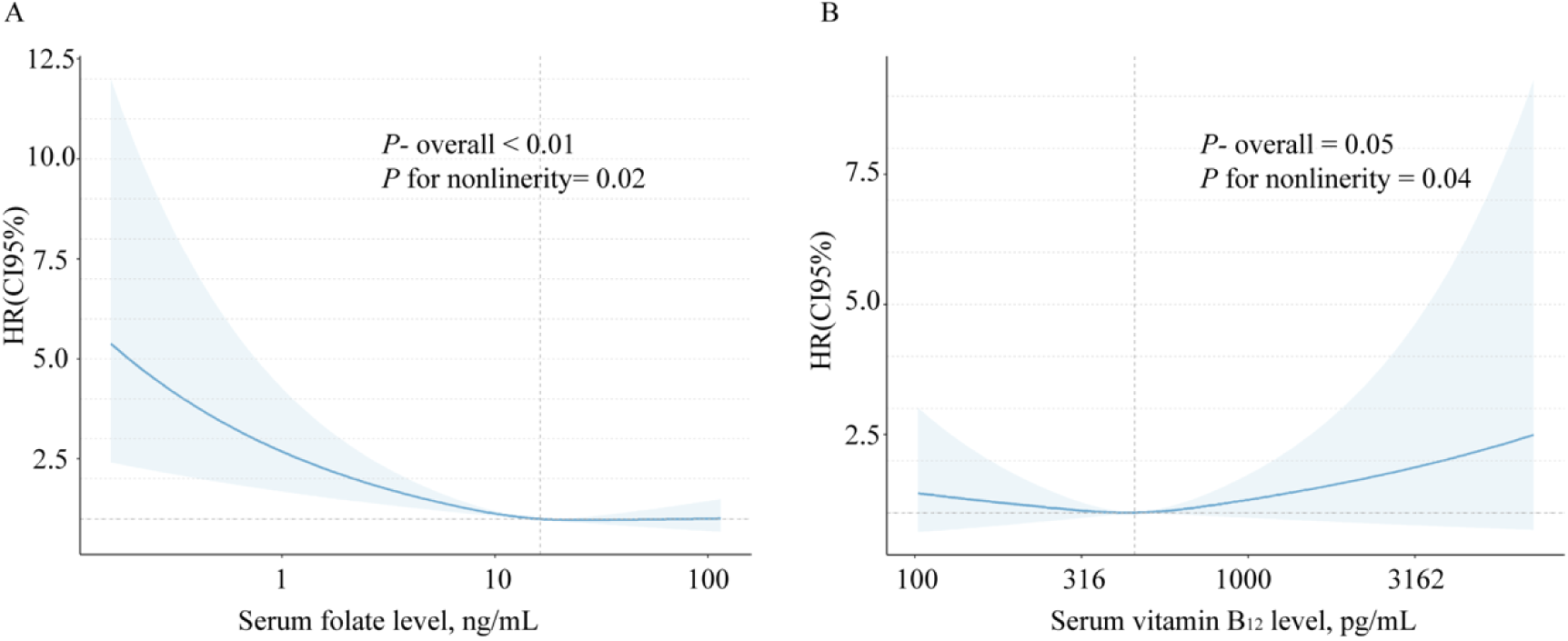
Restricted cubic spline plots of the associations between serum folate, vitamin B_12_ levels and cardiovascular mortality among non-diabetic adults. Hazard ratios (solid lines) and 95% CIs (shaded areas) were adjusted for age, gender, race, education level, PIR, smoking status, alcohol consumption, physical activity, HEI-2015, glycemic status, hypertension, hypercholesterolemia, hypertensive drug use, hyperlipidemia drug use, heart failure, CHD, angina, AMI, stroke, BMI, eGFR, TG, TC, LDL-C, HDL-C, and NHANES cycles. Abbreviations: AMI, acute myocardial infarction; BMI, body mass index; CHD, coronary heart disease; CI: confidence interval; eGFR, estimated glomerular filtration rate; HEI, healthy eating index; HDL-C, high-density lipoprotein cholesterol; HR: hazard ratio; LDL-C, low-density lipoprotein cholesterol; NHANES, National Health and Nutrition Examination Survey; TC, total cholesterol; TG, triglyceride; PIR, poverty-to-income ratio.

**Table 3.** Threshold effect analysis of serum folate and vitamin B_12_ levels on cardiovascular mortality.

| Serum folate level, ng/mL | HR | 95% CI | *P* value |
| --- | --- | --- | --- |
| Cox regression | 0.64 | 0.46, 0.87 | <0.01** |
| Piecewise binary logistic regression models |  |  |  |
| folate < 19.5ng/mL | 0.45 | 0.29, 0.70 | <0.01** |
| folate ≥ 19.5ng/mL | 0.85 | 0.38, 1.88 | 0.69 |
| *P* for log likelihood ratio test |  |  | <0.01** |
| Serum vitamin B<sub>12</sub> level, pg/mL | HR | 95%CI | *P* value |
| Cox regression | 1.25 | 0.85, 1.82 | 0.25 |
| Piecewise binary logistic regression models |  |  |  |
| vitamin B<sub>12</sub> < 436.52pg/mL | 0.70 | 0.23, 2.10 | 0.52 |
| vitamin B<sub>12</sub> ≥ 436.52pg/mL | 1.84 | 1.09, 3.11 | 0.02* |
| *P* for log likelihood ratio test |  |  | <0.01** |

### 3.3 Serum vitamin B_12_ level and cardiovascular mortality

The multivariate logistic regression analysis showed that, compared with the reference group (the first quartile), serum vitamin B_12_ levels in the second, third and fourth quartiles were not significantly associated with the risk of cardiovascular mortality in any of the models (**Table 2**).

The RCS analysis revealed a significant non-linear association between serum vitamin B_12_ levels and cardiovascular mortality (*P* for nonlinearity = 0.04) (**Figure 2B**). The two-piecewise Cox regression models identified that the inflection point of serum vitamin B_12_ level was 436.52 pg/mL. When serum vitamin B_12_ level was ≥ 436.52 pg/mL, each 1-unit increase in serum vitamin B_12_ level was associated with an 84% increased risk of cardiovascular mortality (HR, 1.84; 95% CI, 1.09-3.11; *P* for log-likelihood ratio < 0.01). However, no significant association between serum vitamin B_12_ levels and cardiovascular mortality was observed below the inflection point (HR, 0.70; 95% CI, 0.23-2.10) (**Table 3**).

### 3.4 Stratified and sensitivity analyses

To further evaluate the impact of serum folate and vitamin B_12_ levels on cardiovascular mortality risk, stratified analyses were performed after full adjustment based on age, gender, race, education level, PIR, smoking status, alcohol consumption, BMI, HEI-2015 score, physical activity, glycemic status, self-reported CVD, hypertension, hypercholesterolemia, and eGFR. Compared to non-smokers and individuals with eGFR < 90 mL/min/1.73m^2^, the inverse associations between serum folate levels and cardiovascular mortality were pronounced among smokers (HR: 0.52; 95% CI: 0.33-0.84; *P* for interaction = 0.02) and individuals with eGFR ≥ 90 mL/min/1.73m^2^ (HR: 0.44; 95% CI: 0.25-0.75; *P* for interaction = 0.01). No significant interactions were observed in other subgroups (**Figure 3**).

**Figure 3.**
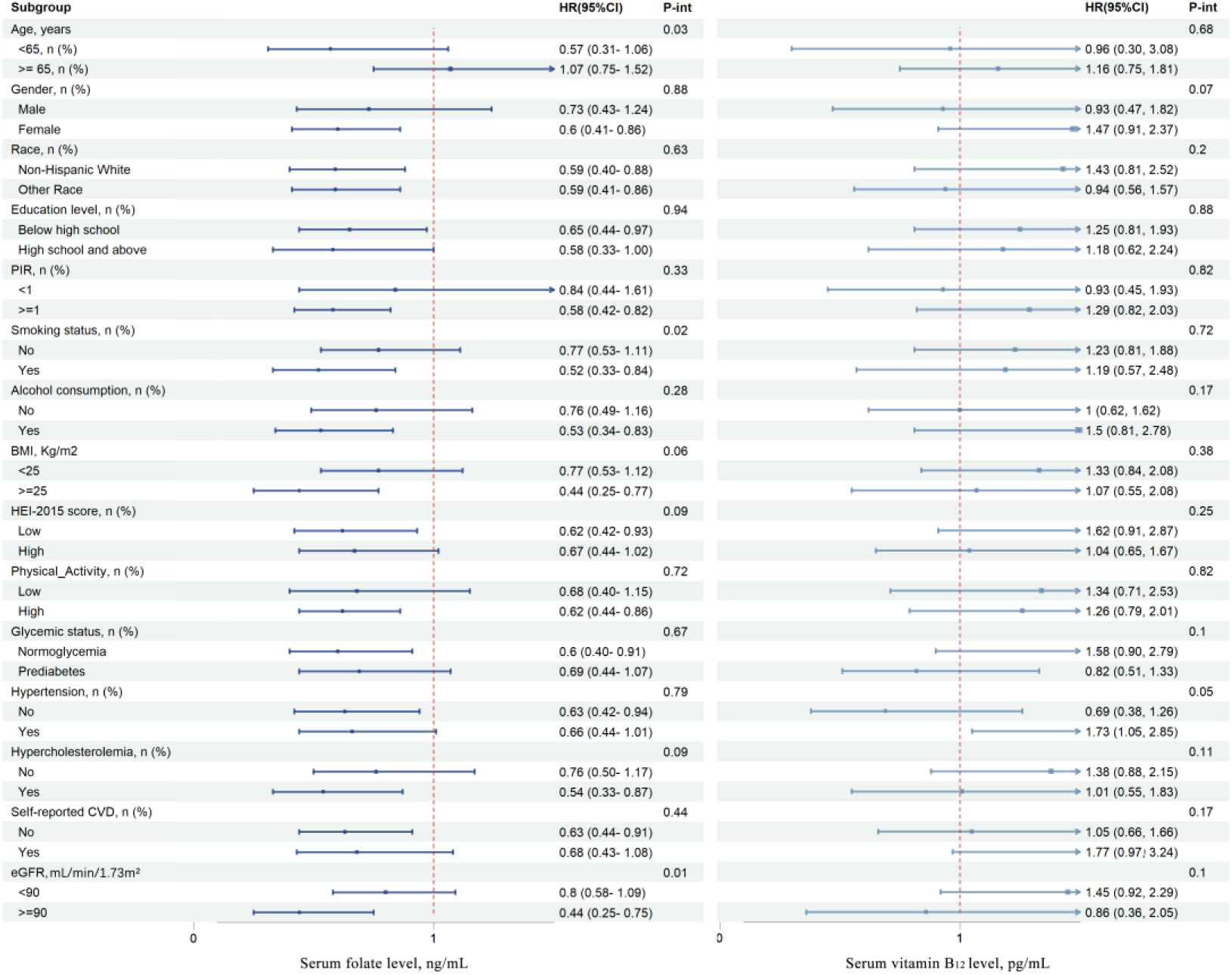
Subgroup and interaction analyses for the associations between serum folate, vitamin B_12_ levels and cardiovascular mortality among adults with non-diabetes. Analyses were adjusted for age, gender, race, education level, PIR, smoking status, alcohol consumption, physical activity, HEI-2015, glycemic status, hypertension, hypercholesterolemia, hypertensive drug use, hyperlipidemia drug use, heart failure, CHD, angina, AMI, stroke, BMI, eGFR, TG, TC, LDL-C, HDL-C, and NHANES cycles. Abbreviations: AMI, acute myocardial infarction; BMI, body mass index; CHD, coronary heart disease; CI: confidence interval; eGFR, estimated glomerular filtration rate; HEI, healthy eating index; HDL-C, high-density lipoprotein cholesterol; HR: hazard ratio; LDL-C, low-density lipoprotein cholesterol; NHANES, National Health and Nutrition Examination Survey; TC, total cholesterol; TG, triglyceride; PIR, poverty-to-income ratio.

In sensitivity analyses, after excluding participants with a history of CVD at baseline, the associations of serum folate and vitamin B_12_ with cardiovascular mortality did not change materially (**Table 1 in Supplement 1**). When participants who died within the first 2 years of follow-up were further excluded, the second and third quartiles of serum folate levels remained significantly associated with cardiovascular mortality when compared to the lowest quartile (**Table 2 in Supplement 1**). After additional adjustment for circulating homocysteine levels, compared to the lowest quartile, the second and third quartiles of serum folate levels were consistently associated with cardiovascular mortality, and the highest quartile of serum vitamin B_12_ levels were also significantly associated with cardiovascular mortality (**Table 3 in Supplement 1**).

## 4. Disccussion

This study is the first to investigate the associations between serum folate, vitamin B_12_ levels, and cardiovascular mortality in a large, nationally representative cohort of non-diabetic US adults. Our findings revealed significant non-linear associations of serum folate and vitamin B12 levels with cardiovascular mortality. Serum folate levels below 19.50 ng/mL and serum vitamin B_12_ levels at or above 436.52 pg/mL were significantly associated with an increased risk of cardiovascular mortality. Stratified and sensitivity analyses validated the robustness of these associations.

Although several previous epidemiological studies have shown an inverse association between folate and CVD risk,^19–21^ the findings remain inconsistent.^22–24^ A Japanese collaborative cohort study identified an inverse relationship between dietary folate intake and heart failure mortality in men, and total cardiovascular mortality in women.^9^ Likewise, a European multicenter study documented that low circulating folate levels were significantly associated with an increased risk of atherosclerosis.^25^ However, a Cochrane review of 8 randomized clinical trials concluded that folate supplementation had no significant impact on MI risk.^24^ These inconsistencies may stem from variations in study design, particularly as the Cochrane review predominantly included individuals with pre-existing CVD or those at high risk. Differences in study endpoints, adjustments for confounding factors, and the inability of supplementation trials to account for dynamic changes in serum folate levels may also contribute to the observed discrepancies. Moreover, population-based studies have reported variable associations between folate and cardiovascular outcomes. The B-PROOF study found no significant effect of folate supplementation on overall CHD incidence in elderly populations.^26^ Among diabetic particiants from the NHANES (1999-2014) and NHANES III (1988-1994), low serum folate levels were linked to a higher risk of cardiovascular mortality.^11^ Whereas in hypertensive populations from NHANES (1999-1994), both low and high serum folate levels were associated with increased cardiovascular mortality.^27^ In the present study, which included 19,402 U.S. non-diabetic adults, serum folate levels below 19.5 ng/mL were significantly associated with an elevated risk of cardiovascular mortality, indicating that the association between folate and CVD risk may vary across different populations.

In contrast, research on the association between vitamin B_12_ and CVD is relatively limited. Several large prospective studies have found no significant effect of vitamin B_12_ supplements on CVD outcomes.^28–30^ First, they used fixed-dose vitamin B_12_ supplements rather than measuring serum vitamin B_12_ levels, which may not accurately reflect its metabolic status. Second, one study enrolled only patients aged 55 and older with pre-existing CVD or diabetes,^28^ while another primarily focused on women aged 42 and older with a history of CVD or high CVD risk.^29^ The specific disease conditions of these populations may have confounded the potential impact of vitamin B_12_ on cardiovascular risk. Furthermore, differences in follow-up durations may have been insufficient to capture the cumulative effects of long-term risk. A study using NHANES data from 1999-2000 through 2013-2014 identified a U-shaped relationship between serum vitamin B_12_ levels and cardiovascular mortality in the general population.^31^ Similarly, analyses of the diabetic population in NHANES (1999-2014) and NHANES III (1988-1994) revealed that both low and high serum vitamin B_12_ levels were associated with an increased risk of cardiovascular mortality.^11^ In this study, we present novel evidence that serum vitamin B_12_ levels exceeding 436.52 pg/mL are significantly associated with an elevated risk of cardiovascular mortality in the non-diabetic population, but no such association was observed for levels below this threshold. This finding suggests that the association between serum vitamin B_12_ and cardiovascular mortality may be population-specific, potentially differing according to metabolic status.

Folate and vitamin B_12_ are essential cofactors in homocysteine metabolism, and numerous epidemiological studies have identified homocysteine as a risk factor for CVD.^5,6^ The association between low serum folate levels and increased CVD risk has often been attributed to the inverse relationship between serum folate and homocysteine levels.^7,32^ However, in our study, the association between low serum folate and increased cardiovascular mortality persisted even after adjusting for homocysteine, indicating the presence of additional mechanisms beyond homocysteine. Previous research has shown that folate supplementation can improve myocardial perfusion in regions affected by diastolic dysfunction, independent of its homocysteine-lowering effect.^33^ In addition, folate has been shown to improve vascular function in patients with CAD by enhancing nitric oxide-mediated, endothelium-dependent vasodilation and reducing vascular superoxide production. Disruptions in folate metabolism have also been associated with various metabolic disorders, including metabolic syndrome, insulin resistance, non-alcoholic fatty liver disease, and altered lipoprotein profiles.^34,35^ Folate deficiency has also been implicated in weight gain and obesity, all of which are recognized risk factors for atherosclerosis and CVD.^36^ Thus, the association between low serum folate levels and cardiovascular mortality may involve multiple overlapping mechanisms.

Conversely, preclinical and clinical studies suggeste that vitamin B_12_ deficiency may negatively affect lipid metabolism, potentially leading to elevated levels of LDL-C, total cholesterol, and subsequent insulin resistance.^37–39^ Despite the association between low B_12_ status and metabolic changes has been documented, our study did not observe a significant association between low vitamin B_12_ levels and cardiovascular mortality. Instead, elevated serum vitamin B_12_ levels were associated with an increased risk of cardiovascular mortality. This paradoxical finding may be attributed to underlying inflammatory conditions, which are known contributors to atherosclerosis and cardiovascular events.^40^ Furthermore, elevated serum vitamin B_12_ levels have been reported in liver diseases, such as hepatitis, cirrhosis, and liver failure.^41^ Potential mechanisms may include upregulated synthesis of haptocorrin and enhanced release of cellular cobalamin, which may coexist with metabolic derangements and elevated cardiovascular risk.^42^ In summary, while the exact mechanisms by which folate and vitamin B_12_ influence cardiovascular outcomes remain unclear, our findings underscore the need for further research to unravel these complex interactions.

## 5. Strengths and limitations

This study has several notable strengths. It is the first to examine the associations between serum folate and vitamin B_12_ levels with cardiovascular mortality specifically in a large, non-diabetic US adult population. The use of a nationally representative sample and thorough adjustments for traditional cardiovascular risk factors enhances the credibility of our findings. Moreover, the extensive stratified and sensitivity analyses further support the consistency and robustness of the observed associations. However, certain limitations should be acknowledged. First, as an observational study, causality cannot be inferred. Second, serum folate and vitamin B_12_ levels were only measured at baseline, which does not account for changes over time. Third, residual confounding may exist due to unmeasured factors, such as dietary intake and genetic variations that affect vitamin metabolism. Finally, the generalizability of our findings to other populations is limited, especially given different dietary habits and health conditions. Future research should validate these associations in diverse cohorts and explore the underlying mechanisms in greater detail.

## 6. Conclusions

In conclusion, our study demonstrates significant non-linear associations of serum folate and vitamin B_12_ levels with cardiovascular mortality in non-diabetic US adults. Low serum folate levels and high vitamin B_12_ levels were both linked to increased risk of cardiovascular mortality. These findings suggest that optimal levels of folate and vitamin B_12_ may play a critical role in cardiovascular health. Future research should aim to clarify underlying mechanisms and inform clinical recommendations.

## Nonstandard Abbreviations and Acronyms

HEI-2015: healthy eating index-2015
HHcy: hyperhomocysteinemia
RCS: restricted cubic spline

## Data Availability

The datasets used during the current study are available from the corresponding author on reasonable request.

## Acknowledgements

We extend our gratitude to all the workers who participated in this study. The authors’ responsibilities were as follows - ZC, DC, XW: designed and conducted the study; ZC: performed statistical analysis, wrote the first draft of the manuscript; DC: interpreted the findings, critically revised the manuscript and provided intellectual content; XW: supervised the study, had primary responsibility for final content. All authors approved the final manuscript.

